# Does the Diabetes Alliance Program Plus Reduce Hospitalisations in Patients with Type 2 Diabetes Attending Primary Care Practices? A Target Trial Protocol for Emulating a Cluster Trial Using Linked General Practice and Tertiary Health Data

**DOI:** 10.1101/2025.05.13.25327569

**Authors:** Joshua Aaron Dizon, Daniel Barker, Shamasunder Acharya, Christopher Oldmeadow, Patrick Skippen, Alexis Hure

## Abstract

**Background:** Effective management of type 2 diabetes is an increasing challenge for health services globally. Integrated care, targeted at increasing capacity in primary care for earlier intervention in type 2 diabetes, may reduce adverse health outcomes, with benefits accrued over time. This protocol aims to describe a causal approach for the evaluation of a specialist-led integrated model of care delivered in Australian general practices, using linked administrative health data.

**Methods:** This protocol outlines an observational cohort study using the Lumos data asset (New South Wales Health), linking general practice and hospital data. It follows the target trial framework, emulating a parallel cluster-randomised trial, and applies the estimands framework proposed by the International Council for Harmonisation of Technical Requirements for Pharmaceuticals for Human Use. It describes the data structure and statistical analysis plan required for causal inference. The primary causal estimand is the effect of the Diabetes Alliance Program Plus, an integrated care intervention, on all-cause hospitalisation rates for adults actively attending general practice with current or newly diagnosed type 2 diabetes over a 5-year follow-up period.

**Discussion:** This protocol applies the target trial framework to emulate a parallel cluster-randomized trial of the Diabetes Alliance Program Plus using linked administrative data. This approach addresses confounding and selection bias inherent in observational evaluations of practice-level interventions and will generate robust real-world evidence to inform policy regarding type 2 diabetes management in primary care settings.

**Trial registration:** ACTRN12622001438741; 10th November 2022, retrospectively registered: https://www.anzctr.org.au/ACTRN12622001438741.aspx.

## INTRODUCTION

Diabetes is a major global health challenge, contributing significantly to disability, premature death, and economic burden across all regions(1). In Australia, approximately 1.3 million of people were living with type 2 diabetes mellitus (T2DM) in 2021, with higher prevalence in remote and socioeconomically disadvantaged areas(2). It is estimated that up to 500000 remain undiagnosed, increasing the risk of complications and premature mortality(3).

T2DM is associated with a wide range of long-term health complications including cardiovascular disease, chronic kidney disease, retinopathy, neuropathy, and psychological distress(4–6). These complications reduce quality of life, increase demand on health services(2), and increase healthcare costs(7). However, early diagnosis and appropriate escalations in treatment can prevent or delay these outcomes.

Despite the central role of primary care in diabetes management in Australia, an estimated 37% of patients do not receive appropriate care for diabetes from general practitioners(8). Barriers include limited access to decision-support tools, inadequate clinical information systems, and insufficient opportunities for continuing professional education(9). These challenges underscore the need for system-level interventions that strengthen primary care capability and capacity, through integrated care pathways, improved digital infrastructure, and stronger linkages with specialist and community services.

One such initiative is the Diabetes Alliance Program Plus (DAP+), an integrated care model originally launched as the Diabetes Alliance Program in 2015 as a partnership between the Hunter New England Local Health District and Hunter New England and Central Coast Primary Health Network(10). The program expanded to DAP+ in 2023 to include the University of Newcastle and Hunter Medical Research Institute. DAP+ aims to build general practice capability and capacity in the Hunter New England Local Health District, New South Wales through a quality-improvement program including: (i) whole-practice data analysis and feedback; (ii) specialist-led case conferencing; and (iii) structured clinician education(10,11).

Early evaluations of DAP+ demonstrated significant short-term improvements in glycaemic control (HbA1c), systolic blood pressure, and cardiovascular risk among participating patients, and the program was well received by both providers and patients(10,12,13). These findings suggest that DAP+ enhances the quality and consistency of diabetes care in general practice by supporting data-driven monitoring, multidisciplinary collaboration, and proactive case management. Improvements in these intermediate markers are clinically meaningful, yet the durability of these effects and their translation into long-term outcomes remain uncertain. Sustained improvements through continued health service engagement could reduce diabetes-related complications, hospitalisations, and premature mortality, with corresponding benefits for health system efficiency and patient wellbeing.

Although randomised controlled trials (RCTs) are the gold standard for assessing intervention effectiveness, they are often impractical in real-world settings. DAP+ was not designed as an RCT and has since been widely implemented as routine practice across the Hunter New England Local Health District. Therefore, this study will apply the target trial emulation framework(14), which enables causal inference from observational data.

This protocol outlines an emulated trial to be conducted using the Lumos data asset (NSW Health) to evaluate the long-term effectiveness of DAP+ on health service use and health outcomes. Specifically, it aims to estimate the causal effect of the DAP+ cluster intervention on all-cause hospital admissions (primary outcome), emergency department presentations, hospital length of stay, lower-limb loss, adherence to testing guidelines, and biomarker levels. It will also determine the feasibility of using the Lumos data asset for evaluating pragmatic, system-level interventions delivered in primary care.

## METHODS AND ANALYSIS

### Target trial emulation

Target trial emulation applies the design principles of RCTs to observational data to strengthen causal inference and reduce bias. The protocol of a target trial specifies eligibility criteria, treatment strategies, assignment procedures, follow-up, outcomes, and the statistical analysis plan. It also defines the causal contrast of interest, typically representing the observational analogues of intention-to-treat and per-protocol effects. In this study, the target trial framework is used to emulate a cluster trial using the Lumos data asset, with all elements of trial design, including eligibility, treatment strategy, assignment, follow-up, outcomes, causal contrast, and analysis, defined *a priori* to minimise bias and enhance reproducibility.

### Data sources and structure

The Lumos data asset is an ethically approved data linkage program led by the New South Wales Ministry of Health, in partnership with Primary Health Networks and the Centre for Health Record Linkage (CheReL)(15). Lumos securely links general practice data with multiple New South Wales (NSW) health data collections, including hospital admission, emergency department, ambulance, non-admitted patient and mortality records, which enables a comprehensive view of patients’ health care access and outcomes. Participating practices provide informed consent for patient records to be linked with statewide health service data, which is enabled through a waiver of individual patient consent under the Lumos Governance Framework(16). All patient and practice level data are deidentified.

The Lumos dataset undergoes regular data quality assessments to ensure completeness, accuracy, and consistency across contributing general practices and linked health system datasets. Previous evaluations have shown the Lumos cohort to be broadly representative of the NSW population with respect to key demographic and clinical characteristics(17).

Use of the Lumos data asset is ethically approved to support public health programs in NSW, including chronic disease management and early intervention. Authorised collaborators access data through the cloud-based Secure Analytics Health Environment (SAPHE) platform. Ethical use of Lumos data is restricted to planning, funding, management, and evaluation of health services(16).

### Target trial specification

A comparative summary of the ideal randomised trial and its emulated target trial is presented in **Table 1**

**Table 1:** Summary of the design features of the target trial protocol for the Diabetes Alliance Program Plus (DAP+) using the Lumos data asset.

| Protocol component | Target trial specification | Target trial emulation |
| --- | --- | --- |
| **Eligibility criteria - Practices** | General practices that consent to participate in the trial and are eligible for randomization to receive the DAP+ intervention or continue usual care. | General practices that consent to data sharing with Lumos.<br>Intervention practices: General practices that consent to receive and have received the DAP+.<br>Control practices: General practices that do not receive the DAP+. |
| **Eligibility criteria - Patients** | Adults ($\geq 18$ years) with a diagnosis of T2DM.<br>Actively attend general practices, defined as $\geq 3$ visits to general practices in the past 2 years.<br>Exclusion criteria:<br>Pregnant women<br>Diabetes diagnoses other than T2DM. | Eligible patients will be identified from eligible practices in the Lumos data asset.<br>Pregnancy status which cannot be directly identified in Lumos data. Consequently, exclusion by pregnancy status cannot be performed. |
| **Treatment strategies** | 1. Receive DAP+ integrated care intervention<br>2. Usual care.<br>Participants continue with their assigned treatment exposure during follow-up. | Same |
| **Assignment procedures** | Practices will be randomly assigned to receive DAP+ (intervention) or continue with usual care. Practices will not be blinded to the intervention. | Randomisation will be emulated via propensity score methods, conditional on covariates identified as important confounders via causal directed acyclic graphs.<br>Matching procedure:<br>Cluster level: many-to-one matching of control to intervention practices, accounting for calendar time, aggregated patient rurality, aggregated patient socioeconomic factors, and high-servicing status<br>One-to-one matching of patients without replacement within matched clusters, accounting for calendar time, age, sex, age at first recorded T2DM diagnosis, rurality, socioeconomic status, comorbidities, frequency of GP |
|  |  | visits, and prior hospitalisations. |
| **Follow-up period** | Follow-up begins at the date following the first case conferencing in each site and continues until death, loss-to-follow-up, or administrative end of follow-up (5 years from baseline), whichever occurs first. | Rolling enrolment with calendar-time matching: treated patients matched to controls with index dates within $\pm 28$ days. Treated index date is the first general practice visit after practice intervention was initiation; control index date is an eligible general practice visit at a non-intervention practice within the matching window. Follow-up begins at index date and continues until death or administrative end of follow-up (5 years from index), whichever occurs first. |
| **Outcomes** | Outcomes are assessed at the individual level; at 1, 3, and 5 years from first exposure to DAP+:<br>• Total all-cause hospitalisations<br>• Total all-cause hospital length of stay<br>• Total potentially preventable hospitalisations<br>• Total T2DM-specific hospitalisation<br>• Total ED presentations<br>• Total potentially avoidable ED presentations<br>• Total T2DM-specific ED presentations<br>• Proportion with T2DM-related lower limb loss<br>• Mortality<br>• HbA1c %, lipids markers (LDL, HDL, triglycerides, and total cholesterol), uACR, and eGFR.<br>• Adherence to testing frequency guidelines for HbA1c, lipids, and kidney function tests. | Same, noting that Urine Albumin-to-Creatinine Ratio (uACR) is not consistently available in Lumos. |
| **Causal contrasts** | Intention-to-treat: effect of assignment to DAP+ versus usual care; | Same |
| **Estimands** | ITT: Sample average treatment effect on the treated among eligible individuals visiting DAP+ practices (effect of at least one exposure to DAP+ enhanced care) | Same |
| **Analysis plan** | Summary of patient baseline characteristics and covariate balance diagnostics among matched cohort.<br>Hierarchical Bayesian models:<br>• Poisson, negative binomial, or | Same |
|  | zero-inflated Poisson regression models for total all-cause hospitalisations, total all-cause hospital length of stay, total potentially preventable hospitalisations, total T2DM-specific hospitalisations, total ED presentations, total potentially avoidable ED presentations, and total T2DM-specific ED presentations.<br>• Logistic regression model for T2DM-related lower limb amputations<br>• Pooled logistic regression model for survival<br>• Logistic regression model for adherence to testing frequency guidelines for HbA1c, lipids, and kidney function tests.<br>• Normal/log-normal regression models for HbA1c, lipids, and uACR, and/or appropriate categorical regression models (logistic, multinomial, or ordinal) for relevant clinical thresholds |  |

### Population and eligibility

The target population includes people with a clinical diagnosis of T2DM who attend general practices participating in the Lumos data linkage program, within NSW. Eligibility is determined dynamically. Patients aged 18 years or older with a current T2DM diagnosis and an active patient status are eligible. Patients not meeting these criteria at a given time may become eligible once the criteria are satisfied. The eligibility definition captures both prevalent and incident T2DM diagnoses within the Lumos data asset.

Baseline covariates will include demographic, socioeconomic, and comorbidity measures defined at the index date, and frequency of visits to general practices, and prior hospitalisations up to two years preceding the index date. Patients must have complete baseline data on all variables required for propensity score estimation.

### Recruitment and follow-up period

Recruitment to the target trial will begin on 1 January 2015 and continue until one year prior to the latest hospital admission record date in the Admitted Patient Data Collection. This ensures that all eligible individuals contribute at least one year of potential follow-up within the five-year outcome window.

Eligible patients from DAP+ intervention practices will enter the cohort on the date of their first eligible visit (index date) after participating in the program. Propensity-matched controls will be selected from eligible patients attending non-DAP+ practices outside the Hunter New England Local Health District, to minimise intervention contamination when practitioners change practices. Outcomes will be evaluated for three fixed intervals relative to the index date: baseline to 1 year, years 2–3, and years 4–5.

Follow-up will begin at the index date and continue until the earliest of (i) five-years post-index, (ii) death (ascertained from the Registry of Births, Deaths, and Marriages), or (iii) administrative censoring at the data cut-off date. Baseline variables for demographic, socioeconomic, and comorbidities will be measured at the index date, and health service access variables within the preceding two years of the index date.

### Treatment assignment procedure

The treatment is DAP+ intervention, delivered in general practice as per the original clinical trial registration (ACTRN12622001438741). The comparator is standard general practice care. Exposure to treatment is defined as the first attendance at a DAP+ practice after meeting eligibility. Exposure to standard care is defined as attendance at a non-DAP+ practice after meeting eligibility.

Because practices are not randomly assigned, treatment assignment will be emulated using propensity score matching to balance measured confounders between exposure groups. The resulting matched cohort will approximate randomisation conditional on observed covariates. Details of the propensity score estimation, matching specification, and balance diagnostics are described in the *Primary Analyses* section.

### Outcomes

The primary outcome is total all-cause hospitalisations, defined as the number of unique patient episodes of care. All-cause hospitalisations will be identified in from the New South Wales Admitted Patient Data Collection (APDC). Multiple same-day admissions will be counted as one episode of care to obtain unique per patient per day events. Totals will be separately calculated for each follow-up interval (baseline to 1 year, years 2-3, and years 4-5).

Secondary outcomes include:

i. hospital length of stay: number of days from admission start date to discharge date as recorded in the APDC. When multiple admissions occur on the same date, the episode with the longest length of stay is used.
ii. potentially preventable hospitalisations: defined according to Australian Institute of Health and Welfare indicators for selected potentially preventable hospitalisation (Table S4 in Additional file 1), with the same definition of episode of care as applied to the primary outcome.
iii. T2DM-specific hospitalisations, defined as a hospitalisation with T2DM diagnosis recorded as the principal diagnosis in the APDC. ICD-10 clinical codes for determining T2DM diagnoses is available in Table S1 in Additional file 1.
iv. ED presentations: identified using the Emergency Department Data Collection (EDDC), with the same definition of episode of care as applied to the primary outcome.
v. potentially avoidable ED presentations: defined according to the Australian Institute of Health and Welfare indicators for selected potentially avoidable GP-type presentations to emergency departments (Table S5 in Additional file 1), with the same definition of episode of care as applied to the primary outcome.
vi. T2DM-specific ED presentations, defined as ED presentations with T2DM diagnosis recorded as the principal diagnosis in the EDDC. The EDDC records diagnosis codes using the ICD-9, ICD-10, or SNOMED-CT code sets. Codes used to identify T2DM-specific ED presentations in each system are listed in Tables S2 and S3 in Additional file 1.
vii. T2DM-related lower-limb loss: defined according to the Australian Institute of Health and Welfare indicator for Hospitalisation for lower limb amputation with T2DM as principal or additional diagnosis (Table S6 in Additional file 1).
viii. all-cause mortality: death dates will be obtained from Registry of Births, Deaths, and Marriages data set.
ix. Adherence to testing frequency guidelines for T2DM pathology monitoring: testing frequency will use definitions from the Royal Australian College of General Practitioners’ annual cycle of care guidelines for T2DM(18). Separate adherence measures will be constructed for HbA1c, lipid profile, and renal function testing. For HbA1c, each test initiates a 6-month coverage window extending from test date. For lipid profile and renal function tests, each test initiates a 12-month coverage window. If a subsequent test occurs within an active coverage window, the coverage is extended from the new test date for the relevant duration. Coverage from tests conducted before the start of a follow-up interval is carried forward into the interval. Coverage intervals that extend beyond the end of a follow-up interval will be truncated at the interval’s end. For each test type, the proportion of months within the interval covered by any active window is calculated per participant. Participants with more than 90 percent coverage for a given test type are classified as adherent to that guideline.
x. T2DM-related biomarker levels (HbA1c, lipid profile, eGFR): Laboratory measurements will be obtained from General Practice Electronic Health Records. For each biomarker, the mean value will be calculated separately for each patient within each follow-up interval. When multiple tests occur within an interval, all available results will contribute to the per patient interval mean. Analyses will be performed separately for each biomarker. Additionally, biomarkers levels may be classified according to clinically relevant thresholds (e.g., HbA1C <7%, 7-8.9%, ≥9% etc.).

### Clarifying the estimands

An estimand defines the treatment effect that a study aims to quantify. Clear specification of estimands clarifies the research question and guides appropriate methodological choices.

Estimands in this protocol are defined according to the International Council for Harmonisation (ICH) E9(R1) guidelines and the framework proposed by Kahan (2024)(19) . The matched population will be used to estimate the intention-to-treat (ITT) effect. Handling of intercurrent events, which are post-baseline events that may affect interpretation, will also be specified. The corresponding estimands are summarised in Table 2.

**Table 2:** Description of the estimands for testing the effectiveness of the Diabetes Alliance Program Plus (DAP+) on hospitalisations compared to standard care in general practice.

| Attribute | Specification |
| --- | --- |
| Estimand | Sample average treatment effect on the treated for the intention-to-treat (ITT) population: effect of at least one exposure to DAP+ intervention (i.e. enhanced care) |
| Population | Adults ($\geq 18$ years) with existing or newly diagnosed T2DM recorded in Lumos, who are active patients of general practices in New South Wales, Australia. |
| Treatment conditions | Intervention: Attendance at a DAP+ enhanced care practice.<br>Comparator: Attendance at a non-DAP+ practice. |
| Endpoint | Primary:<br>Total number of hospitalisations, counted as unique admissions per day, summed over three prespecified time intervals (baseline-year 1, years 2-3, and years 4-5). The primary estimand targets the treatment effect on the total number of hospitalisations per participant within these intervals, evaluated using a longitudinal model with time and treatment-by-time interaction terms.<br>Secondary:<br>Total hospital length of stay*, total potentially preventable hospitalisations, total T2DM-specific hospitalisations, total emergency department presentations, total potentially avoidable emergency department presentations, total T2DM-specific ED presentations, T2DM-related lower limb loss, adherence to T2DM-related pathology monitoring guidelines, and T2DM-related biomarker levels, each assessed over the three prespecified time intervals (baseline-year 1, years 2-3, years 4-5). The estimand for each is the treatment effect on the mean value (total count, proportion, or continuous measure as appropriate) within intervals.<br>All-cause mortality, analysed as a discretised time-to-event outcome with survival status evaluated at 1, 3, and 5 years. The estimand is the treatment effect on the probability of survival at each prespecified time point. |
| Summary Measure | Count outcomes:<br>Incidence rate ratio comparing treatment vs control within time intervals.<br>Binary outcomes:<br>Risk ratio comparing treatment vs control within time intervals.<br>Continuous outcomes<br>Mean difference between treatment and control within time intervals.<br>Discretised time-to-event outcomes:<br>Risk difference of survival at 1, 3, and 5 years. |
| Handling of<br>intercurrent<br>events | Loss of active attendance: Treatment policy strategy. Analyses will use exposure assigned at baseline, regardless of whether patients become inactive during follow-up.<br>Treatment switching: Treatment policy strategy. Analyses will use exposure assigned at baseline, regardless of post-baseline switching between DAP+ and non-DAP+ practices.<br>Death during follow-up: While-alive strategy. For all outcomes except all-cause mortality, data will be included from index date until the date of death. |

*\*When multiple admissions occur on the same date, the episode with the longest length of stay is used*

### Primary Analyses

### General principles

All analyses will comply with Lumos data governance requirements, including suppression of cells with counts ≤5, which will be reported as ‘≤5’.

### Causal analysis principles

An observational analogue of the ITT effect will be applied to the primary analysis. DAP+ exposure will be compared to the relevant comparator (standard care) using a propensity matched cohort.

Relevant covariates for creating the propensity matched cohort were identified using the directed acyclic graph (DAG) in Figure 1, which was informed by literature review(20–23) and clinical domain knowledge. It was determined that treatment assignment is predominantly affected by geographic factors and general practice characteristics.

**Figure 1.**
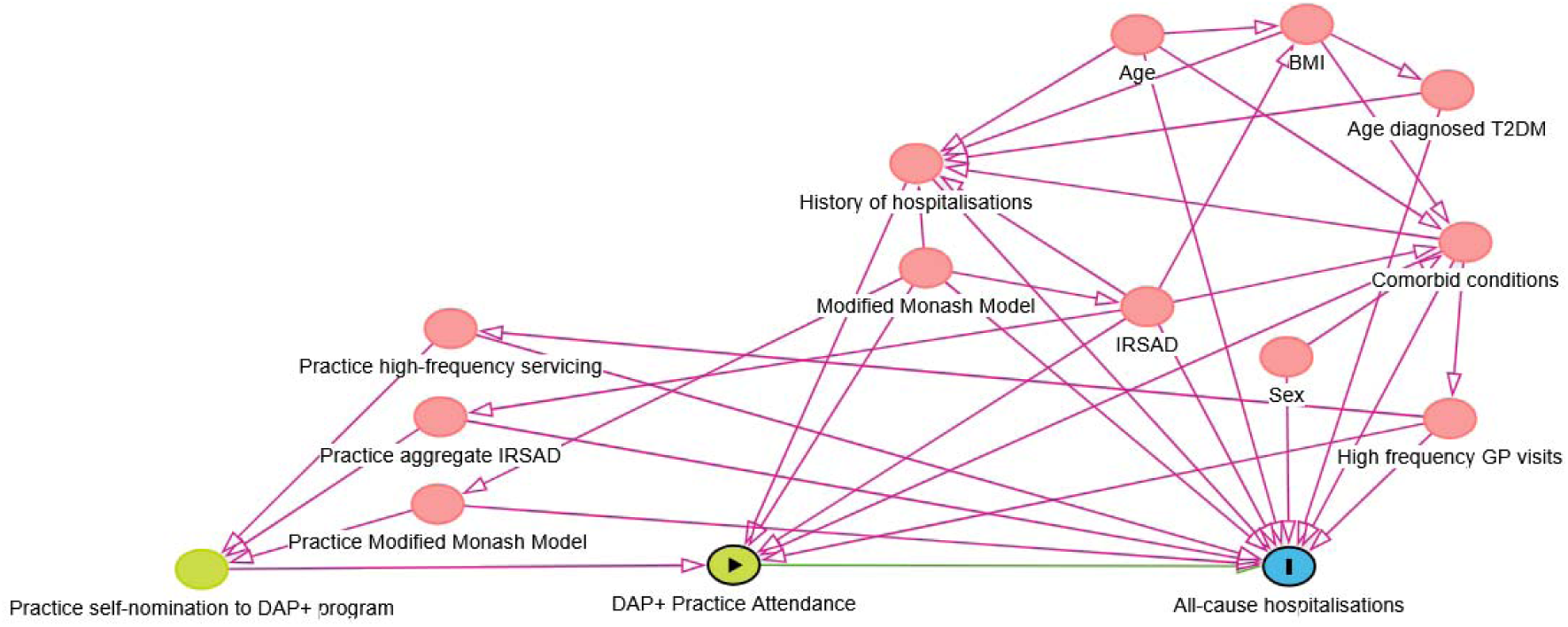
Directed Acyclic Graph representing relations between DAP+ exposure, outcome, and confounders

Access to the DAP+ enhanced care intervention depends not only on individual characteristics but also on whether a patient’s general practice self-nominated to participate in DAP+. Individual exposure can occur only within participating practices. Self-nomination may be influenced by the implementation of DAP+, which primarily targets, though is not limited to, regional general practices(10). Practice-level aggregates of Index of Relative Socioeconomic Advantage and Disadvantage (IRSAD, an Australian national measure ranking areas by relative socioeconomic conditions based on income, education, and employment)(24), geographic classification under the Australian Modified Monash Model (MMM, which categorises areas by remoteness and population size)(25), and operational features such as high-frequency servicing (>30% of patients attending ≥12 times over two years)(20) were identified as key determinants of participation. Because these factors are also associated with health outcomes, they form cluster-level biasing pathways upstream of the exposure–outcome relationship.

At the individual level, factors influencing general practice attendance include sociodemographic status via IRSAD, geographic remoteness via MMM, comorbidities, frequency of general practice visits, and prior hospitalisations. In Australia, multimorbidity is more prevalent in regional and remote areas than urban settings(26). As these factors are associated with both health outcomes and the likelihood of attending practices in different socioeconomic or rural settings, they may also form biasing pathways between the intervention and outcomes. Although BMI, age, and sex lie on the causal pathway, they are not required in the minimal adjustment set. Adjustment for individual MMM, IRSAD, and comorbid conditions is sufficient to close the identified individual-level biasing paths in the DAG.

### Descriptive statistics

Summary statistics of baseline characteristics will be stratified by treatment group and will include where available in the Lumos data asset; the age at study entry, sex, BMI, IRSAD, MMM, comorbidities, and health service access indicators. Summary measures will be mean and standard deviation (SD) for continuous variables, and frequencies and percentages for categorical variables.

### Balancing cluster and individual confounders via propensity scores

The minimal adjustment set required to identify the causal effect of receiving the intervention includes practice-level MMM, aggregate IRSAD, high-frequency servicing indicator, and individual-level MMM, IRSAD, comorbidities, and health service use indicators. To improve the precision of estimates, individual BMI, sex, and age will also be included as additional covariates.

Before individual-level matching, intervention and control practices will be exactly matched on MMM classification, aggregate IRSAD, and high-frequency servicing status. Each intervention cluster may be paired with one or more control clusters to balance cluster-level confounding on these variables.

After balancing cluster-level confounders by forming cluster-similar groups, intervention-exposed individuals will be matched to control individuals within these groups. Individual matching will be based on propensity scores estimated using a logistic regression model, with individual exposure to the intervention at baseline as the outcome and individual level confounders as predictors. Each DAP+ participant will be matched to the control with the closest propensity score who did not attend a DAP+ practice at baseline. Matches will have propensity scores within 0.2 pooled standard deviations of the logit of the propensity score to improve covariate balance between matched pairs(27). Matching will be performed one-to-one without replacement, and unmatched individuals will be excluded.

Covariate balance will be assessed in terms of standardised mean differences after propensity matching to determine whether balancing was successful. Interactions of variables and non-linear functional forms of continuous variables will be considered to iteratively improve covariate balance, as necessary.

### Outcome analysis

All outcomes will be analysed using longitudinal Bayesian hierarchical models to account for clustering and repeated measurements within individuals and dyad pairs. Models will be fit using the propensity-matched sample. All models will include fixed effects for treatment, time, and their interaction, and random intercepts to account for repeated measures within clusters and dyad pairs. Weakly informative priors will be specified for all fixed and random effects to improve model regularisation and ensure stable Markov chain Monte Carlo sampling without imposing restrictive assumptions.

### Primary outcome

The primary outcome is the total number of hospitalisations in all time intervals. For each patient, all hospitalisations within an interval will be summed to obtain a single patient-level total. This outcome will be modelled using a Bayesian hierarchical Poisson, negative-binomial, or zero-inflated Poisson model, as selected by model fit, evidence of overdispersion, or excess zeroes. The model will estimate incidence rate ratios (IRR) with 95% credible intervals (CrI).

### Secondary outcomes

Secondary outcomes include total hospital length of stay, total potentially preventable hospitalisations, total T2DM-specific hospitalisations, total ED presentations, total potentially avoidable ED presentations, and T2DM-specific ED presentations in all time intervals. Each will be analysed using the same modelling considerations as the primary outcome to estimate IRR with 95% CrI. Statewide data indicate low event rates for T2DM-specific hospitalisations (3.7%)(2), raising the possibility of nonconvergent models in the matched cohort. If models fail to converge, results will be reported descriptively by treatment group and time interval. Similar considerations apply to T2DM-specific ED presentations.

T2DM-related lower limb loss will be analysed using a logistic regression model. Posterior predictions of event probabilities will be used to estimate relative risks (RR) with 95% CrI.

All-cause mortality will be analysed using a pooled logistic regression model for discretised time-to-event. Cumulative survival probabilities will be obtained via posterior predictions to estimate risk differences (RD) with 95% CrI at 1, 3, and 5 years.

Pathology biomarker outcomes will be analysed using normal or log-normal regression models based on empirical distribution of each biomarker. For biomarkers with clinically informative thresholds, secondary analyses will be conducted using logistic, multinomial, or ordinal regression models where appropriate.

Compliance with annual cycle-of-care testing for metabolic markers will be analysed using logistic regression models.

## DISCUSSION

Evidence indicates that the risk of hospital admission among patients with type 2 diabetes mellitus (T2DM) is associated with elevated HbA1c levels(28,29) and other routinely measured primary care markers, including increased uACR, elevated systolic blood pressure, and reduced high-density lipoprotein (HDL) cholesterol(29). The association between elevated HbA1c and macrovascular and microvascular complications has been well established since 2000, with each 1% HbA1c reduction associated with 21% lower diabetes-related endpoints and 37% lower microvascular complication (30),underscoring the importance of metabolic control in preventing hospitalisations and other adverse outcomes. In the pilot evaluation of DAP+, improvements in diabetes-related metabolic markers, particularly HbA1c, were observed among T2DM patients attending DAP+-enabled general practices(10,11). Given the established link between these markers and diabetes-related complications, these findings suggest that DAP+ may contribute to improved long-term outcomes for patients with T2DM through enhanced metabolic control. However, as the pilot study was not designed to assess long-term endpoints, the effect of DAP+ on longer-term outcomes remains an important question warranting further investigation. Potentially preventable hospitalisations are widely used to monitor the performance of primary health care systems and to evaluate the effectiveness of primary care interventions in Australia and internationally(31). As the DAP+ program aims to strengthen chronic disease management in general practice, reductions in potentially preventable hospitalisation rates represent a plausible downstream indicator of its effectiveness in improving diabetes care and long-term health outcomes.

Ideally, the effectiveness of an intervention would be assessed using a RCT. However, as the DAP+ program has been progressively implemented across the Hunter and New England region since 2015, conducting a new RCT in this setting is not feasible due to potential contamination and spillover from prior program exposure, and the cost of running such a large trial. Implementing an RCT in other health districts would also pose substantial practical and resource challenges, as it would require establishing the DAP+ model of care de novo within a different health service context. There are also ethical considerations, given the existing evidence supporting its intermediate benefits (REFS), making it questionable to withhold access to enhanced care. Furthermore, in regions without the DAP+ model, patients have limited access to metropolitan specialist care and remain at increased risk of premature and progressive diabetes-related complications due to undertreated disease.

DAP+ has been progressively implemented across the Hunter New England Local Health District in partnership with the Hunter New England and Central Coast Primary Health Network since 2015, and administrative health data from participating general practices are routinely captured through the Lumos data linkage program in New South Wales(17). This linkage provides a rich longitudinal dataset suitable for evaluating the long-term effects of DAP+. Using these existing data within a target trial emulation framework(14) offers a practical and robust alternative to conducting a new RCT. Previous studies have shown that well-designed emulated trials using observational data can yield treatment effect estimates consistent with those obtained from true RCTs(32).

Although this protocol employs propensity score methods to balance key confounders, residual confounding cannot be fully eliminated and remains a limitation of any observational study(33–36). Unmeasured differences may exist across Local Health Districts and Primary Health Networks in which practices are nested, potentially influencing diabetes management. However, all operate under New South Wales Health governance with uniform policy, funding, and clinical frameworks. Matching within geographic strata (metropolitan, regional, rural, remote) further limits systematic region-level bias.

Lumos participation is voluntary among general practices, introducing potential selection bias in the data collections. As of November 2025, more that 7 million people across NSW are represented with approximately one-third of all general practices having consented to data-sharing across the state (Add Ref). The 2023 Lumos evaluation reported 91% demographic representativeness to the broader New South Wales population(37), providing reasonable assurance that the Lumos data set captures statewide demographic diversity.

While recognising the limitations inherent to observational analyses, this study will generate the best available evidence to inform general practice-centred models of care for type 2 diabetes mellitus in rural and remote communities in NSW.

## DECLARATIONS

### Ethics approval and consent to participate

The Diabetes Alliance Program Plus has ethics approval from the Hunter New England Human Research Ethics Committee (2024/ETH01649), registered with the University of Newcastle (R-2024-0073). The Lumos data asset has ethics approval from the Population Health Services Research Ethics Committee (2019/ETH00660) with a waiver of individual participant consent for approved secondary data uses.

## Consent for publication

Not applicable.

## Availability of data and materials

No new datasets were generated in this study.

## Authors’ contributions

JAD, DB, CO and AH contributed to the statistical analysis plan and statistical rationale described in this article. AH and SA provided clinical rationale for this protocol. All authors were involved in the discussions and editing of the manuscript and provide approval for publication.

## Patient and public involvement

Patients and/or the public were not involved in the design, conduct, reporting, or dissemination plans of this research.

## Acknowledgements

The authors acknowledge the Lumos program, a partnership of the NSW Ministry of Health and NSW Primary Health Networks made possible through the participation of general practices across NSW. Record linkage was carried out by the Centre for Health Record Linkage.

## REFERENCES

1. Ong KL, Stafford LK, McLaughlin SA, Boyko EJ, Vollset SE, Smith AE, et al. Global, regional, and national burden of diabetes from 1990 to 2021, with projections of prevalence to 2050: a systematic analysis for the Global Burden of Disease Study 2021. The Lancet. 2023 Jul 15;402(10397):203–34.

2. Australian Institute of Health and Welfare [Internet]. [cited 2025 Nov 11]. Diabetes: Australian Facts. Available from: https://www.aihw.gov.au/reports/diabetes/diabetes/contents/summary

3. Diabetes Australia [Internet]. [cited 2025 Nov 11]. Diabetes in Australia. Available from: https://www.diabetesaustralia.com.au/about-diabetes/diabetes-in-australia/

4. Galiero R, Caturano A, Vetrano E, Beccia D, Brin C, Alfano M, et al. Peripheral Neuropathy in Diabetes Mellitus: Pathogenetic Mechanisms and Diagnostic Options. Int J Mol Sci. 2023 Feb 10;24(4).

5. Dal Canto E, Ceriello A, Rydén L, Ferrini M, Hansen TB, Schnell O, et al. Diabetes as a cardiovascular risk factor: An overview of global trends of macro and micro vascular complications. Eur J Prev Cardiol. 2019 Dec;26(2_suppl):25–32.

6. Akshatha S, Nayak UB. The psychological distress associated with type 2 diabetes mellitus represents an unmet need for drug discovery. Medicine in Drug Discovery. 2024;23:100196.

7. Lee CMY, Colagiuri R, Magliano DJ, Cameron AJ, Shaw J, Zimmet P, et al. The cost of diabetes in adults in Australia. Diabetes Research and Clinical Practice. 2013 Mar;99(3):385–90.

8. Runciman WB, Hunt TD, Hannaford NA, Hibbert PD, Westbrook JI, Coiera EW, et al. CareTrack: assessing the appropriateness of health care delivery in Australia. Medical Journal of Australia. 2012 Jul;197(2):100–5.

9. Brown JB, Harris SB, Webster-Bogaert S, Wetmore S, Faulds C, Stewart M. The role of patient, physician and systemic factors in the management of type 2 diabetes mellitus. Family Practice. 2002 Aug 1;19(4):344–9.

10. Acharya S, Philcox AN, Parsons M, Suthers B, Luu J, Lynch M, et al. Hunter and New England Diabetes Alliance: innovative and integrated diabetes care delivery in general practice. Australian Journal of Primary Health. 2019;25(3):219.

11. Acharya S, Taylor R, Parsons M, Attia J, Leigh L, Oldmeadow C, et al. Spillover effects from a type 2 diabetes integrated model of care in 22,706 Australians: an open cohort stepped wedge trial. BMC Endocr Disord. 2024 Sep 10;24(1):183.

12. Harris ML, Kuzulugil D, Parsons M, Byles J, Acharya S. “They were all together … discussing the best options for me”: Integrating specialist diabetes care with primary care in Australia. Health & Social Care in the Community. 2021 Sep;29(5):e135–43.

13. Taylor RM, Acharya SH, Parsons ME, Ranasinghe UP, Kuzulugil DO, Fleming KC, et al. Australian practice nurses’ perspectives on integrating specialist diabetes care with primary care: a qualitative study. Fam Pract. 2025 Apr 12;42(3):cmaf020.

14. Hernán MA, Robins JM. Using Big Data to Emulate a Target Trial When a Randomized Trial Is Not Available: Table 1. American Journal of Epidemiology. 2016 Apr 15;183(8):758–64.

15. NSW Health [Internet]. [cited 2025 Nov 11]. About Lumos. Available from: https://www.health.nsw.gov.au/lumos/Pages/about.aspx

16. NSW Health [Internet]. 2025 [cited 2025 Nov 11]. Lumos Data Governance Framework. Available from: https://www.health.nsw.gov.au/lumos/Publications/data-governance-framework.pdf

17. Correll P, Feyer AM, Phan PT, Drake B, Jammal W, Irvine K, et al. Lumos: a statewide linkage programme in Australia integrating general practice data to guide system redesign. Integrated Healthcare Journal. 2021 May 17;3(1):e000074.

18. The Royal Australia College of General Practitioners [Internet]. 2020. Management of type 2 diabetes: A handbook for general practice.

19. Kahan BC, Hindley J, Edwards M, Cro S, Morris TP. The estimands framework: a primer on the ICH E9(R1) addendum. BMJ. 2024 Jan 23;384:e076316.

20. NSW Health [Internet]. 2021 [cited 2025 Nov 11]. General practice activity can affect hospital visits. Available from: https://www.health.nsw.gov.au/lumos/Factsheets/gp-activity-insight.pdf

21. Phillips A. Health status differentials across rural and remote Australia. Australian Journal of Rural Health. 2009 Feb;17(1):2–9.

22. Britt HC, Harrison CM, Miller GC, Knox SA. Prevalence and patterns of multimorbidity in Australia. Medical Journal of Australia. 2008 Jul;189(2):72–7.

23. Glenister KM, Guymer J, Bourke L, Simmons D. Characteristics of patients who access zero, one or multiple general practices and reasons for their choices: a study in regional Australia. BMC Family Practice. 2021 Dec;22(1):2.

24. Australian Bureau of Statistics [Internet]. [cited 2025 Nov 11]. Socio-Economic Indexes for Areas (SEIFA), Australia. Available from: https://www.abs.gov.au/statistics/people/people-and-communities/socio-economic-indexes-areas-seifa-australia/latest-release#index-of-relative-socio-economic-advantage-and-disadvantage-irsad-

25. Versace VL, Skinner TC, Bourke L, Harvey P, Barnett T. National analysis of the Modified Monash Model, population distribution and a socio[economic index to inform rural health workforce planning. Australian Journal of Rural Health. 2021 Oct;29(5):801–10.

26. Australian Institute of Health and Welfare [Internet]. [cited 2025 Nov 11]. Multimorbidity in Australia. Available from: https://www.aihw.gov.au/reports/chronic-disease/multimorbidity-in-australia/contents/how-common-is-multimorbidity

27. Austin PC. An Introduction to Propensity Score Methods for Reducing the Effects of Confounding in Observational Studies. Multivariate Behav Res. 2011 May;46(3):399–424.

28. Bo S, Ciccone G, Grassi G, Gancia R, Rosato R, Merletti F, et al. Patients with type 2 diabetes had higher rates of hospitalization than the general population. Journal of Clinical Epidemiology. 2004 Nov;57(11):1196–201.

29. Tomlin AM, Dovey SM, Tilyard MW. Risk factors for hospitalization due to diabetes complications. Diabetes Research and Clinical Practice. 2008 May;80(2):244–52.

30. Stratton IM, Adler AI, Neil HA, Matthews DR, Manley SE, Cull CA, et al. Association of glycaemia with macrovascular and microvascular complications of type 2 diabetes (UKPDS 35): prospective observational study. BMJ. 2000 Aug 12;321(7258):405–12.

31. Australian Commission on Safety and Quality in Health Care [Internet]. 2017 [cited 2025 Nov 11]. A guide to the potentially preventable hospitalisations indicator in Australia. Available from: https://www.safetyandquality.gov.au/publications-and-resources/resource-library/guide-potentially-preventable-hospitalisations-indicator-australia

32. Wang SV, Schneeweiss S, RCT-DUPLICATE Initiative, Franklin JM, Desai RJ, Feldman W, et al. Emulation of Randomized Clinical Trials With Nonrandomized Database Analyses: Results of 32 Clinical Trials. JAMA. 2023 Apr 25;329(16):1376.

33. Fewell Z, Davey Smith G, Sterne JAC. The Impact of Residual and Unmeasured Confounding in Epidemiologic Studies: A Simulation Study. American Journal of Epidemiology. 2007 Jun 24;166(6):646–55.

34. Howards PP. An overview of confounding. Part 1: the concept and how to address it. Acta Obstetricia et Gynecologica Scandinavica. 2018 Apr;97(4):394–9.

35. Verbeek JH, Whaley P, Morgan RL, Taylor KW, Rooney AA, Schwingshackl L, et al. An approach to quantifying the potential importance of residual confounding in systematic reviews of observational studies: A GRADE concept paper. Environment International. 2021 Dec;157:106868.

36. Arah OA. Bias Analysis for Uncontrolled Confounding in the Health Sciences. Annual Review of Public Health. 2017 Mar 20;38(1):23–38.

37. NSW Health [Internet]. 2023 [cited 2025 Nov 11]. Lumos Evaluation Report 3. Available from: https://www.health.nsw.gov.au/lumos/Publications/lumos-evaluation-report-3.pdf

