## Additional file 1 for "Does the Diabetes Alliance Program Plus Reduce Hospitalisations in Patients with Type 2 Diabetes Attending Primary Care Practices? A Target Trial Protocol for Emulating a Cluster Trial Using Linked General Practice and Tertiary Health Data"

### SUPPLEMENTARY MATERIAL

Table S1. Mapping of key variables within Lumos

| Criterion | Data table/s | Variables | Method of constructing criterion |
| --- | --- | --- | --- |
| Patient identifier | GPEHR.*, APDC.*, EDDC.*, RBMD.* | *PPN* | As recorded. |
| Primary Health Network identifier | GPEHR.Patient | *AREA_ID* | As recorded. |
| Practice identifier | GPEHR.Patient | *PRACTICE_ID* | As recorded. |
| Patient demographics (age, sex, IRSAD, Modified Monash Model) | GPEHR.Patient | *SEX, IRSAD_DECILE, MMM_1* | As recorded, except for age.  Age will be calculated as years from *YEAR_OF_BIRTH* to baseline date. |
|  | Master.Summary | *YEAR_OF_BIRTH* |  |
| Comorbidities | GPEHR.PatientChronicDisease | *CLASSIFICATION,*  *DIAGNOSIS_DATE* | Comorbidities as recorded in Lumos will be used. Lumos records 25 unique chronic conditions: *Acute Coronary Syndrome, ADHD, Asthma, Atrial Fibrillation, Autism, Bipolar, Cancer, Carotid Stenosis, Chronic Kidney Disease, Chronic Osteoporosis, Coeliac Disease, COPD, CVD, Depression, Diabetes Type 1, Diabetes Type 2, Gestational Diabetes, Heart Failure, Hyperlipidaemia, Liver Disease, Mental Health Conditions, Myocardial Infarction, Schizophrenia, Stroke, Undefined Diabetes* |
| Eligibility | GPEHR.PatientEncounter | *ENCOUNTER_DATE,* | Patient eligibility will be defined at each unique *ENCOUNTER_DATE*.  Eligibility will be determined if the patient is aged 18 or over, has a current diagnosis of T2DM at the *ENCOUNTER_DATE*, and has at least 3 previous observations recorded in the GPEHR.PatientEncounter with unique *ENCOUNTER_DATES* in the 12 months prior to the current *ENCOUNTER_DATE*.  Current T2DM diagnosis at time of encounter will be defined as having a *CLASSIFICATION* corresponding to T2DM and DIAGNOSIS DATE preceding ENCOUNTER_DATE, or DIAGNOSIS_CODE corresponding to T2DM and *EPISODE_START_DATE* preceding *ENCOUNTER_DATE.* |
|  | GPEHR.Patient | *YEAR_OF_BIRTH* |  |
|  | GPEHR.PatientChronicDisease | *CLASSIFICATION, DIAGNOSIS_DATE* |  |
|  | APDC.PatientDiagnosis  APDC.PatientVisit | *DIAGNOSIS_CODE, EPISODE_START_DATE* |  |
| i. Practice intervention exposure  ii. Patient intervention exposure | GPEHR.PatientEncounter  GPEHR.PatientBilling | *ENCOUNTER_DATE, BILLING_ITEM_NUMBER* | i. Practice intervention exposure will be defined at the general practice level. Any patient encounters at a given practice with an *ENCOUNTER_*DATE that is after DATE_DAP*,* will be defined as having received treatment.  ii. Patient intervention exposure will be defined at the patient level for each encounter. Patients will not be considered exposed until both eligibility and practice intervention exposure coincides. The earliest *ENCOUNTER_DATE* where eligibility is met will be used as the index date. |
|  | DAPPracticeIdentifier^†^ | *PRACTICE_ID,*  *DATE_DAP* |  |
| i. Metabolic marker measurements (HbA1c, cholesterol, HDL, LDL, Triglycerides, EGFR)  ii. Compliance to individualised diabetes assessment intervals*^‡^* | GPEHR.PatientPathology | *DATE, PATHOLOGY_TYPE, VALUE, UNITS* | i. Each row in GPEHR.PatientPathology wherein *PATHOLOGY_TYPE* corresponds to each respective metabolic marker will be used.  ii. Compliance will be determined using *DATE*. For a given calendar period, patients will be deemed compliant from the date of measurement up to the maximum recommended time interval between measurement. If a new measurement occurs within the current interval, compliance is extended up to the maximum interval starting from the date of the new measurement. If no new measurements occur within the maximum interval, patients will be deemed non-compliant until a new measurement occurs. |
| i. Hospitalisations,  ii. length-of-stay of hospitalisation  iii. Potentially preventable hospitalisations.  iv. length-of-stay of potentially preventable hospitalisations  v. T2DM-related lower-limb loss | APDC.PatientVisit | *PPN, EPISODE_START_DATE, EPISODE_END_DATE,*  *APDC_EPISODE_ID* | i. Each record in APDC.PatientVisit represents an episode-of-care. Only one episode of care will be counted per PPN, per EPISODE_START_DATE  ii. Calculated as the number of days from *EPISODE_START_DATE* and *EPISODE_END_DATE* from a hospitalisation episode of care.  iii. *PROCEDURE_CODE* and *PROCEDURE_ BLOCK* from APDC.PatientProcedures and *DIAGNOSIS_CODE* from APDC.PatientDiagnosis will be linked to APDC.PatientVisit via *APDC_EPISODE_IDs*. DIAGNOSIS_CODE and PROCEDURE_BLOCK will be used to flag potentially preventable hospitalisations. Definitions of potentially preventable hospitalisations are presented in Supplementary Table 3.  iv. Calculated as the number of days from *EPISODE_START_DATE* and *EPISODE_END_DATE* from a potentially preventable hospitalisation episode of care.  v. Data set linking similar to iii. to flag hospitalisations with T2DM-related limb loss. Definition of T2DM-related lower limb loss is presented in Supplementary Table 5. |
|  | APDC.PatientDiagnosis | *DIAGNOSIS_CODE, APDC_EPISODE_ID* |  |
|  | APDC.PatientProcedure | *PROCEDURE_DATE, PROCEDURE_CODE,*  *PROCEDURE_BLOCK, APDC_EPISODE_ID* |  |
| i. ED presentations  ii. Potentially avoidable ED presentations | EDDC.PatientVisit | *PPN, ARRIVAL_DATE, ACTUAL_DEPARTURE_DATE* | i. Each record in EDDC.PatientVisit represents an episode-of-care. Only one episode of care will be counted per PPN, per ARRIVAL_DATE  ii. |
| Survival status | RBDM.DeathRegistration | *PPN, DATE_OF_DEATH* | Survival status will be defined using *DATE_OF_DEATH*. |

^†^*The DAP+ practice identifying data set is external to the Lumos data asset.*

Table S2. ICD-9-Clinical Modification and ICD-10-Australian Modification Diabetes mellitus codes

| **ICD-9-Clinical Modification (US and Australian Versions) 1993-94 to 1997-98** | | |
| --- | --- | --- |
| **Diabetes Type** | **Diabetes Complication** | **ICD Code** |
| *Determined by the*  *fifth digit*  Type 1 Diabetes  …………….…. 1 or 3  Type 2 Diabetes  …………….…. 0 or 2 | Diabetes without mention of complication  Diabetes with ketoacidosis  Diabetes with hyperosmolarity  Diabetes with other coma  Diabetes with renal complication  Diabetes with ophthalmic complication  Diabetes with neurological complication  Diabetes with peripheral circulatory disorders  Diabetes with other specified complications  Diabetes with unspecified complication | 250.0_  250.1_  250.2_  250.3_  250.4_  250.5_  250.6_  250.7_  250.8_  250.9_ |
| **ICD-10-Australian Modification First Edition 1998-99 to 1999-2000** | | |
| **Diabetes Type** | **Diabetes Complication** | **ICD Code** |
| *Determined by the prefix*  Type 1 Diabetes  …………….…. E10  Type 2 Diabetes  …………….…. E11  Other specified diabetes  …………….…. E13  Unspecified diabetes  …………….…. E14 | Diabetes with coma | E1_.0_ |
|  | Diabetes with ketoacidosis | E1_.1_ |
|  | Diabetes with renal complications | E1_.2_ |
|  | Diabetes with ophthalmic complications | E1_.3_ |
|  | Diabetes with neurological complications | E1_.4_ |
|  | Diabetes with peripheral circulatory complications | E1_.5_ |
|  | Diabetes with other specified complications | E1_.6_ |
|  | Diabetes with multiple complications | E1_.7_ |
|  | Diabetes with unspecified complications | E1_.8_ |
|  | Diabetes without complications | E1_.9_ |
| **ICD-10-Australian Modification Second Edition 2000-01** | | |
| **Diabetes type** | **Diabetes complication** | **ICD code** |
| *Determined by the prefix*  Type 1 Diabetes …………….…. E10  Type 2 Diabetes  …………….…. E11  Other specified diabetes  …………….…. E13  Unspecified diabetes  …………….…. E14 | Diabetes with ketoacidosis without coma | E1_.11 |
|  | Diabetes with ketoacidosis with coma | E1_.12 |
|  | Diabetes with lactic acidosis without coma | E1_.13 |
|  | Diabetes with lactic acidosis with coma | E1_.14 |
|  | Diabetes with ketoacidosis, with lactic acidosis without coma | E1_.15 |
|  | Diabetes with ketoacidosis, with lactic acidosis with coma | E1_.16 |
|  | Diabetes with renal complication | E1_.2 |
|  | … with renal complication, unspecified | E1_.20 |
|  | … with incipient diabetic nephropathy | E1_.21 |
|  | … with established diabetic nephropathy | E1_.22 |
|  | … with end-stage renal disease | E1_.23 |
|  | … with other specified renal complication | E1_.29 |
|  | *Diabetes with ophthalmic complication* | E1_.3 |
|  | … with ophthalmic complication, unspecified | E1_.30 |
|  | … with background retinopathy | E1_.31 |
|  | … with preproliferative retinopathy | E1_.32 |
|  | … with proliferative retinopathy | E1_.33 |
|  | … with other retinopathy | E1_.34 |
|  | … with advanced ophthalmic disease | E1_.35 |
|  | … with diabetic cataract | E1_.36 |
|  | … with other specified complication | E1_.39 |
|  | *Diabetes with neurological complication* | E1_.4 |
|  | … with neuropathy, unspecified | E1_.40 |
|  | … with diabetic mononeuropathy | E1_.41 |
|  | … with diabetic polyneuropathy | E1_.42 |
|  | … with diabetic autonomic neuropathy | E1_.43 |
|  | … with other specified neurological complication | E1_.49 |
|  | *Diabetes with circulatory complication* | E1_.5 |
|  | … with circulatory complication, unspecified | E1_.50 |
|  | … with peripheral angiopathy, without gangrene | E1_.51 |
|  | … with peripheral angiopathy, with gangrene | E1_.52 |
|  | … with other specified circulatory complication | E1_.59 |
|  | *Diabetes with other specified complication* | E1_.6 |
|  | … with diabetic musculoskeletal & connective tissue complication | E1_.61 |
|  | … with skin and subcutaneous tissue complication | E1_.62 |
|  | … with periodontal complication | E1_.63 |
|  | … with hypoglycaemia | E1_.64 |
|  | … with other specified complication | E1_.69 |
|  | *Diabetes with multiple complications* | E1_.7 |
|  | … with multiple microvascular complications | E1_.71 |
|  | … with foot ulcer due to multiple sources | E1_.73 |
|  | Diabetes with unspecified complications | E1_.8 |
|  | Diabetes without complication | E1_.9 |

Australian Institute of Health and Welfare [Internet]. [cited 2025 Nov 14]. The impact of ICD coding standard changes for diabetes hospital morbidity data. Available from: https://www.aihw.gov.au/getmedia/85711ef4-0ecd-4a29-a92e-ee2a39fdced2/iicdcscdhmd.pdf?v=20230605170459&inline=true

Table S3. SNOMED CT-AU Diabetes Mellitus code extract from NTCS Emergency department reference set

| **SNOMED CT-AU** | |
| --- | --- |
| **Code** | **Description** |
| 73211009 | Diabetes mellitus |
| 268519009 | Diabetes mellitus uncontrolled |
| 11530004 | Unstable diabetes mellitus |
| 44054006 | Type 2 diabetes mellitus |
| 43959009 | Cataract due to diabetes mellitus |
| 74627003 | Complication due to diabetes mellitus |
| 237632004 | Hypoglycaemic event due to diabetes |
| 420422005 | Ketoacidosis due to diabetes mellitus |
| 20130731 | Retinopathy due to diabetes mellitus |
| 443694000 | Type 2 diabetes uncontrolled |
| 422126006 | Hyperosmolar coma due to diabetes mellitus |
| 26298008 | Ketoacidotic coma due to diabetes mellitus |
| 190389009 | Type 2 diabetes with ulcer |
| 230572002 | Diabetic neuropathy |
| 25093002 | Disorder of eye due to diabetes mellitus |
| 420756003 | Cataract due to diabetes mellitus type 2 |
| 200687002 | Cellulitis of foot due to diabetes mellitus |
| 127013003 | Disorder of kidney due to diabetes mellitus |
| 368051000119109 | Hyperglycaemia due to type 2 diabetes |
| 421750000 | Ketoacidosis due to type 2 diabetes mellitus |
| 421895002 | Peripheral vascular disorder due to diabetes mellitus |
| 422099009 | Diabetic oculopathy associated with type 2 diabetes mellitus |
| 422034002 | Diabetic retinopathy associated with type 2 diabetes mellitus |
| 310505005 | Hyperosmolar non-ketotic state due to diabetes mellitus |
| 421847006 | Ketoacidotic coma due to type 2 diabetes mellitus |
| 314902007 | Peripheral angiopathy due to type 2 diabetes mellitus |
| 127014009 | Diabetic peripheral angiopathy |
| 237621004 | Diabetic severe hyperglycaemia |
| 422183001 | Diabetic skin ulcer |
| 390834004 | Nonproliferative diabetic retinopathy |
| 421966007 | Non-ketotic non-hyperosmolar coma due to diabetes mellitus |
| 422166005 | Peripheral circulatory disorder due to type 2 diabetes mellitus |
| 371087003 | Diabetic foot ulcer |
| 111556005 | Diabetic ketoacidosis without coma |

*SNOMED CT-AU codes from the NTCS Emergency Department Reference Set used to identify ED presentations where type 2 diabetes mellitus was the principal diagnosis. Codes were identified using CSIRO’s Shrimp browser** *to search term 'diabetes', returning 49 codes, of which 15 were excluded: Gestational diabetes mellitus, Vasopressin-related polyuria, Type 1 diabetes mellitus, Arginine vasopressin resistance, AVP-D - arginine vasopressin deficiency, Primary polydipsia, Partial vasopressin resistance, Unstable type 1 diabetes mellitus, Type 1 diabetes mellitus with ulcer, Gangrene due to type 1 diabetes mellitus, Ketoacidosis due to type 1 diabetes mellitus, Diabetic oculopathy due to type 1 diabetes mellitus, Ketoacidotic coma due to type 1 diabetes mellitus, Peripheral circulatory disorder due to type 1 diabetes mellitus, Diabetic retinopathy associated with type 1 diabetes mellitus. The remaining 34 codes included 11 type 2 diabetes-specific codes and 23 generic diabetes codes; generic codes were attributed to type 2 diabetes given the cohort's confirmed type 2 diabetes diagnosis.
*CSIRO [Internet]. [cited 2025 Nov 14]. Shrimp. Available from: https://ontoserver-wp.csiro.au/our-solutions/shrimp/*

###### Supplementary Table S4. Potentially preventable hospitalisation definitions

| **Category** | **ICD-10-AM** | **Comments** |
| --- | --- | --- |
| ***Vaccine preventable conditions*** | | |
| Pneumonia and influenza (vaccine-preventable) | J10, J11, J13, J14 | In any diagnosis. Exclude people under 2 months. Rehabilitation records are excluded. |
| Other vaccine-preventable conditions | A08.0, A35, A36, A37, A80, B01, B05, B06, B16.1, B16.9, B18.0, B18.1, B26, G00.0 | In any diagnosis. Rotaviral Enteritis (A08.0) included for records with separation date 1 July 2007 onwards. Rehabilitation records are excluded. |
| ***Chronic*** | | |
| Asthma | J45, J46 | As principal diagnosis. Exclude children aged less than 4 years. Rehabilitation records are excluded. |
| Congestive cardiac failure | I50, I11.0, J81 | As principal diagnosis. Exclude cases with the following cardiac procedure codes: Blocks 600-606, 608-650, 653-657, 660-664, 666, 669-682, 684-691, 693, 705-707, 717 and codes 33172-00[715], 33827-01[733], 34800-00[726], 35412-00[11], 38721-01[733], 90217-02[734], 90215-02[732]. Rehabilitation records are excluded. |
| Diabetes complications | E10, E11, E13, E14 | As principal diagnosis. Rehabilitation records are excluded. |
| COPD | J20, J41, J42, J43, J44 | J41-J44 as principal diagnosis. J20 as principal diagnosis with additional diagnoses of J41, J42, J43, J44. Rehabilitation records are excluded. |
| Bronchiectasis | J47, J20 | As principal diagnosis. J20 only with additional diagnosis of J47. Rehabilitation records are excluded. |
| Angina | I20, I24.0, I24.8, I24.9 | As principal diagnosis. Exclude cases according to the list of procedures excluded from the Congestive cardiac failure category above. Rehabilitation records are excluded. |
| Iron deficiency anaemia | D50.1, D50.8, D50.9 | As principal diagnosis. Rehabilitation records are excluded. |
| Hypertension | I10, I11.9 | As principal diagnosis.Exclude cases with procedure codes according to the list of procedures excluded from the Congestive cardiac failure category above. Rehabilitation records are excluded. |
| Nutritional deficiencies | E40, E41, E42, E43, E55.0, E64.3 | As principal diagnosis. Rehabilitation records are excluded. |
| Rheumatic heart diseases | I00, I01, I02, I05, I06, I07, I08, I09 | As principal diagnosis. Rehabilitation records are excluded. |
| ***Acute*** | | |
| Pneumonia (not vaccine-preventable) | J15.3, J15.4, J15.7, J16.0 | In any diagnosis. Exclude people under 2 months. Rehabilitation records are excluded. |
| Urinary tract infections, including pyelonephritis | N10, N11, N12, N13.6, N15.1, N15.9, N28.9, N39.0, N39.9 | As principal diagnosis. Rehabilitation records are excluded. |
| Perforated/bleeding ulcer | K25.0, K25.1, K25.2, K25.4, K25.5, K25.6, K26.0, K26.1, K26.2, K26.4, K26.5, K26.6, K27.0, K27.1, K27.2, K27.4, K27.5, K27.6, K28.0, K28.1, K28.2, K28.4, K28.5, K28.6 | As principal diagnosis. Rehabilitation records are excluded. |
| Cellulitis | L02, L03, L04, L08, L88, L98.0, L98.3 | As principal diagnosis. Exclude cases with any procedure except those in blocks 1820 to 2016, or if procedure is 30216-00, 30216-01, 30216-02, 30676-00, 30223-01, 30223-02, 30064-00, 90660-00, 90661-00, and this is the only listed procedure. Rehabilitation records are excluded. |
| Pelvic inflammatory disease | N70, N73, N74 | As principal diagnosis. Rehabilitation records are excluded. |
| Ear, nose and throat infections | H66, J02, J03, J06, J31.2 | As principal diagnosis. Rehabilitation records are excluded. |
| Dental conditions | K02, K03, K04, K05, K06, K08, K09.8, K09.9, K12, K13, K14.0 | As principal diagnosis. Rehabilitation records are excluded. |
| Convulsions and epilepsy | G40, G41, R56 | As principal diagnosis. Rehabilitation records are excluded. |
| Eclampsia | O15 | As principal diagnosis. Rehabilitation records are excluded. |
| Gangrene | R02, I70.24, E09.52 | R02 in any diagnosis. I70.2 and E09.52 as principal diagnosis. Rehabilitation records are excluded. |

HealthStats NSW [Internet]. Potentially preventable hospitalisations. Available from: https://www.healthstats.nsw.gov.au/page/potentially-preventable-hospitalisation-codes

###### Supplementary Table S5. Potentially avoidable ED presentation definition

| **Computation description:** | Potentially avoidable General Practitioner (GP)-type presentations are defined as presentations to public hospital emergency departments with a Type of visit of *Emergency presentation* where the patient:   - was allocated a triage category of 4 (Semi-urgent: within 60 minutes) or 5 (Non-urgent: within 120 minutes) and - did not arrive by ambulance, or police or correctional vehicle and - was not admitted to the hospital, not referred to another hospital, or did not die. |
| --- | --- |

Australian Institute of Health and Welfare [Internet]. [cited 2025 Nov 14]. National Healthcare Agreement: PI 19–Selected potentially avoidable GP-type presentations to emergency departments, 2021. Available from: https://meteor.aihw.gov.au/content/725791

###### Supplementary Table S6. T2DM-coded lower limb loss definition

| **Definition** | Number of hospital separations for lower limb amputation (ACHI Block 1533, procedure codes:  44370-00, 44373-00, 44367-00, 44367-01, 44367-02), with type 2 diabetes (ICD-10-AM: E11) as a principal diagnosis. |
| --- | --- |

Australian Institute of Health and Welfare [Internet]. 2020. Indicators for the Australian National Diabetes Strategy 2016–2020: data update. Available from: https://www.aihw.gov.au/reports/diabetes/diabetes-indicators-strategy-2016-2020/contents/goal-3-reduce-the-occurrence-of-diabetes-related-c/indicator-3-12-hospitalisation-for-lower-limb-ampu
